# Clinical factors associated with racial differences in the prevalence of occult hypoxemia: a retrospective case-control study

**DOI:** 10.1101/2024.03.28.24305036

**Authors:** Sofia K. Mettler, Nipith Charoenngam, Aunchalee Jaroenlapnopparat, Courtney Tern, Nutchapon Xanthavanij, Sofia Economidou, Matthew J. Strand, Brian D. Hobbs, Matthew Moll, Michael H. Cho

## Abstract

**Background:** Recent studies showed that Black patients more often have falsely normal oxygen saturation on pulse oximetry compared to White patients. However, whether the racial differences in occult hypoxemia are mediated by other clinical differences is unknown.

**Methods:** We conducted a retrospective case-control study utilizing two large ICU databases (eICU and MIMIC-IV). We defined occult hypoxemia as oxygen saturation on pulse oximetry within 92-98% despite oxygen saturation on arterial blood gas below 90%. We assessed associations of commonly measured clinical factors with occult hypoxemia using multivariable logistic regression and conducted mediation analysis of the racial effect.

**Results:** Among 24,641 patients, there were 1,855 occult hypoxemia cases and 23,786 controls. In both datasets, Black patients were more likely to have occult hypoxemia (unadjusted odds ratio 1.66 [95%-CI: 1.41-1.95] in eICU and 2.00 [95%-CI: 1.22-3.14] in MIMIC-IV). In multivariable models, higher respiratory rate, PaCO2 and creatinine as well as lower hemoglobin were associated with increased odds of occult hypoxemia. Differences in the commonly measured clinical markers accounted for 9.2% and 44.4% of the racial effect on occult hypoxemia in eICU and MIMIC-IV, respectively.

**Conclusion:** Clinical differences, in addition to skin tone, might mediate some of the racial differences in occult hypoxemia.

## Introduction

Pulse oximetry is an essential tool to identify hypoxemia. However, recent studies reported that Black patients are more likely to have falsely normal pulse oximetry (SpO2) readings compared to White patients while their arterial oxygen saturation measured by blood gas (SaO2) is abnormally low – a phenomenon referred to as “occult hypoxemia” (1-6). Occult hypoxemia may delay appropriate treatment, resulting in systemic racial disparities in medical care and outcomes (2-6).

While differences in skin pigmentation are suspected to contribute to the racial differences in pulse oximetry accuracy (7, 8), other possible mechanisms have been proposed. These include severity of illness, underlying comorbidities, hemoglobinopathy as well as serum biochemistry that may affect light absorbance (9-12). However, the relative contribution of these factors to the occurrence of occult hypoxemia and its racial difference remains unclear. Using two large intensive care unit (ICU) databases, we aimed to identify factors that are associated with occult hypoxemia and quantify how much of the racial difference in occult hypoxemia is attributable to differences in these factors using mediation analysis.

## Methods

We conducted a retrospective case-control study utilizing two publicly available databases: the electronic Intensive Care Unit collaborative research database (eICU) and Medical Information Mart for Intensive Care (MIMIC-IV). The eICU database contains clinical data of ICU admissions between 2014 and 2015 at more than 200 hospitals across the United States (13). The MIMIC-IV database contains clinical data of ICU admissions at an academic tertiary care center in Boston, Massachusetts between 2008 and 2019 (14). Both databases are deidentified and approved by the institutional review boards of Massachusetts Institute of Technology and participating hospitals. For all our analyses, we used the eICU as our initial dataset, and MIMIC-IV as validation.

### Inclusion criteria

We included adult patients (18 to 89 years-old) with at least one SaO2 measurement. The inclusion flow chart is available in Figure 1. We limited our analyses to Black and White patients when examining the racial effect.

**Figure 1.**
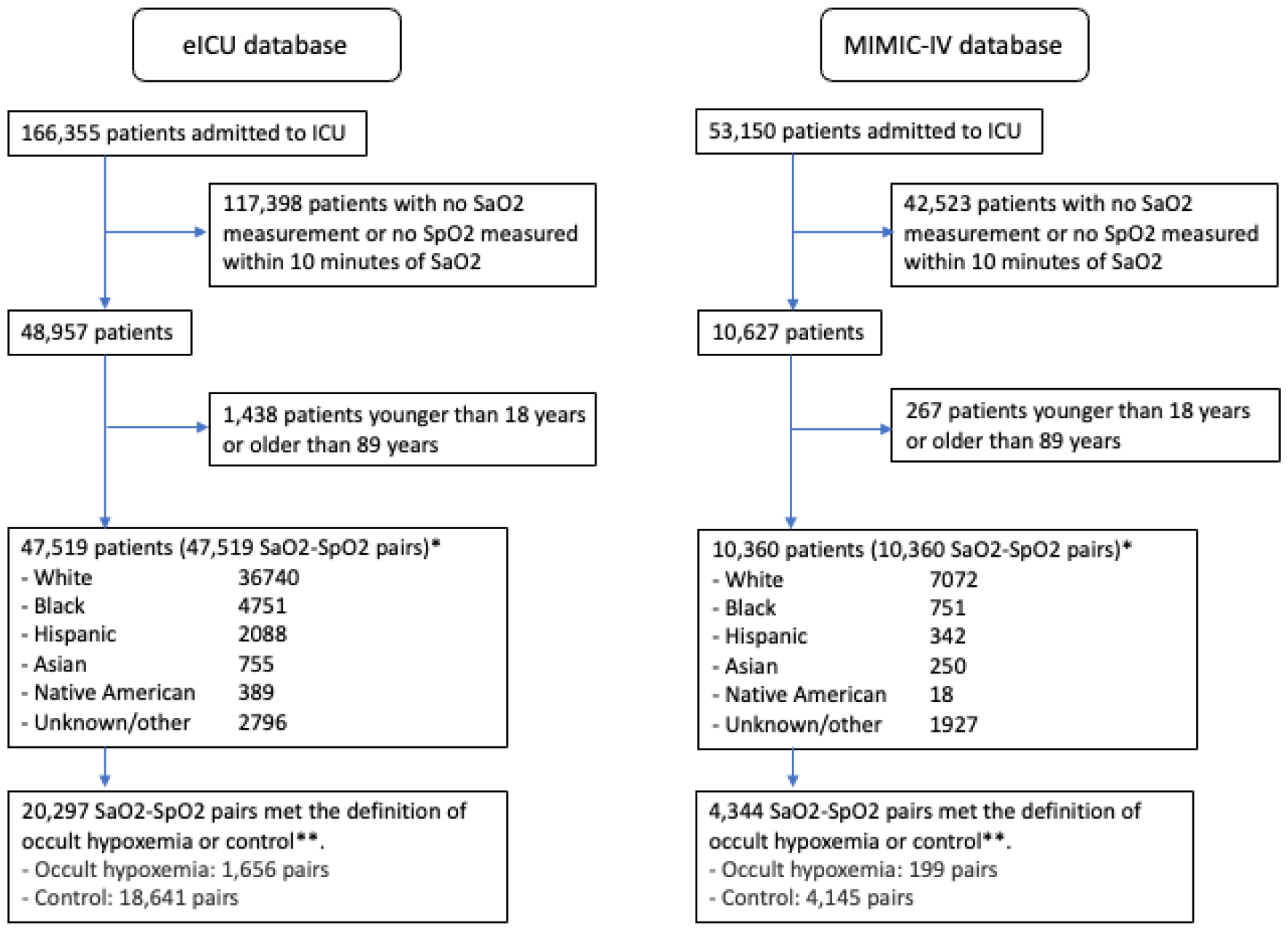
Inclusion flow chart. *One SaO2-SpO2 pair (the earliest) per patient was included. **The condition of defining occult hypoxemia cases or controls was that SpO2 is within 92-98% and SaO2 is 70% or above.

### SaO2-SpO2 pairs

As done in previous studies (2, 6), we included only the first SaO2-SpO2 pair for a given patient to limit potential confounding from repeat measurements. We paired the first SaO2 measurement of each patient with the closest pulse oximetry (SpO2) value, allowing a maximum time difference of 10 minutes between the two. We repeated the analysis allowing a maximum time difference of 5 minutes.

### Definition of occult hypoxemia cases and controls

We defined occult hypoxemia cases as SaO2 70-89% measured by arterial blood gas while SpO2 is between 92% and 98%, because hypoxemia with SaO2 <90% is reported to be associated with worse outcomes (15, 16), and controls as SaO2 ≥ 90% with SpO2 between 92% and 98%.

### Covariate selection

Our criteria for covariate selection were the following: 1) routinely available clinical data, excluding factors that can be affected by external circumstances, such as treatment or diagnosis code, 2) available in both eICU and MIMIC-IV, and 3) not missing in more than 30% of observations. The selected covariates were demographics (age, gender, race, BMI), vital signs (heart rate, respiratory rate, systolic blood pressure and temperature) as well as laboratory tests (PaCO2, arterial pH, hemoglobin, MCV, white blood cell count, platelet count, Na, K, creatinine, and glucose levels). We excluded highly correlated variables in the multivariable models. For example, we included hemoglobin into the model and excluded hematocrit, MCHC and MCH.

### Statistical analysis

We compared the means of continuous variables using two sample t-tests, count data using chi-square tests and proportions using two-sample proportion z-tests. We assessed associations of occult hypoxemia with covariates using multivariable logistic regression models (see Supplement A for the directed acyclic graph considered in our study). We performed mediation analysis and calculated the proportion of the racial effect mediated by covariates (17, 18). We performed all analyses in eICU first and attempted to reproduce the results in MIMIC-IV. Statistical significance was defined as p-value smaller than 0.05. All statistical analyses are performed using the statistical software R (version 4.2.1). Additional methods including data extraction and outlier removal are explained in detail in Supplement B.

## Results

A total of 57,879 SaO2-SpO2 pairs from 57,879 patients were available (47,519 from eICU and 10,360 from MIMIC-IV). Among these, we identified occult hypoxemia cases and controls in 24,641 patients.

### Patient characteristics

Among 20,297 patients in eICU whose SpO2 ranged between 92% and 98%, occult hypoxemia (SaO2<90%) was found in 1,656 patients (8.16%). The overall prevalence of occult hypoxemia was lower in MIMIC-IV (199 patients among 4,344 patients, 4.58%). Patient characteristics are compared between occult hypoxemia cases and controls in Table 1. Many of the statistically significant differences found in eICU were replicable in MIMIC-IV.

**Table 1.**
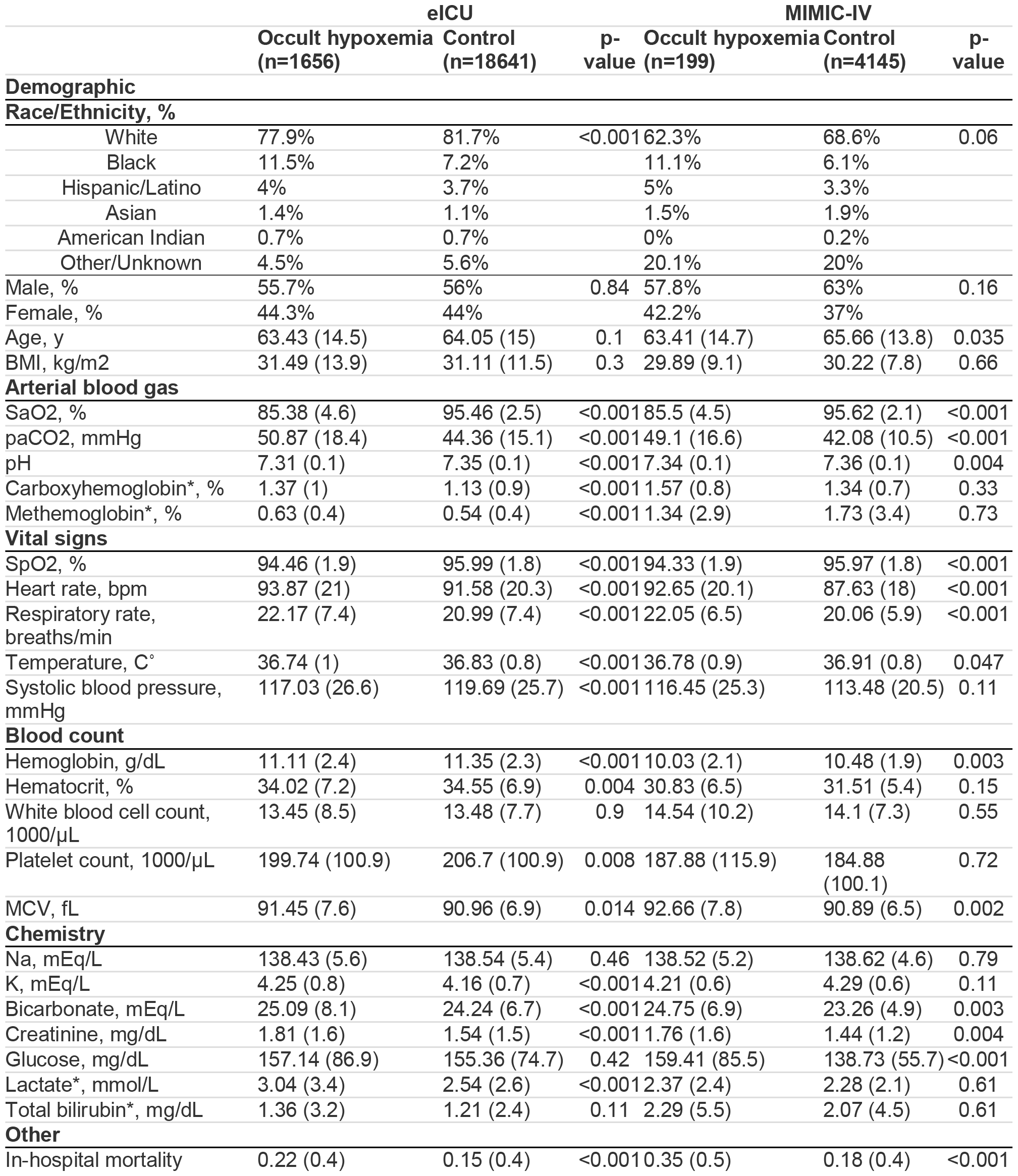
Patient characteristics by occult hypoxemia vs. control. Univariable comparison between cases and controls. *Missing values in more than 30% of observations.

|  | eICU |  |  | MIMIC-IV |  |  |
| --- | --- | --- | --- | --- | --- | --- |
|  | Occult hypoxemia<br>(n=1656) | Control<br>(n=18641) | p-value | Occult hypoxemia<br>(n=199) | Control<br>(n=4145) | p-value |
| **Demographic** |  |  |  |  |  |  |
| **Race/Ethnicity, %** |  |  |  |  |  |  |
| White | 77.9% | 81.7% | <0.001 | 62.3% | 68.6% | 0.06 |
| Black | 11.5% | 7.2% |  | 11.1% | 6.1% |  |
| Hispanic/Latino | 4% | 3.7% |  | 5% | 3.3% |  |
| Asian | 1.4% | 1.1% |  | 1.5% | 1.9% |  |
| American Indian | 0.7% | 0.7% |  | 0% | 0.2% |  |
| Other/Unknown | 4.5% | 5.6% |  | 20.1% | 20% |  |
| Male, % | 55.7% | 56% | 0.84 | 57.8% | 63% | 0.16 |
| Female, % | 44.3% | 44% |  | 42.2% | 37% |  |
| Age, y | 63.43 (14.5) | 64.05 (15) | 0.1 | 63.41 (14.7) | 65.66 (13.8) | 0.035 |
| BMI, kg/m2 | 31.49 (13.9) | 31.11 (11.5) | 0.3 | 29.89 (9.1) | 30.22 (7.8) | 0.66 |
| **Arterial blood gas** |  |  |  |  |  |  |
| SaO2, % | 85.38 (4.6) | 95.46 (2.5) | <0.001 | 85.5 (4.5) | 95.62 (2.1) | <0.001 |
| paCO2, mmHg | 50.87 (18.4) | 44.36 (15.1) | <0.001 | 49.1 (16.6) | 42.08 (10.5) | <0.001 |
| pH | 7.31 (0.1) | 7.35 (0.1) | <0.001 | 7.34 (0.1) | 7.36 (0.1) | 0.004 |
| Carboxyhemoglobin*, % | 1.37 (1) | 1.13 (0.9) | <0.001 | 1.57 (0.8) | 1.34 (0.7) | 0.33 |
| Methemoglobin*, % | 0.63 (0.4) | 0.54 (0.4) | <0.001 | 1.34 (2.9) | 1.73 (3.4) | 0.73 |
| **Vital signs** |  |  |  |  |  |  |
| SpO2, % | 94.46 (1.9) | 95.99 (1.8) | <0.001 | 94.33 (1.9) | 95.97 (1.8) | <0.001 |
| Heart rate, bpm | 93.87 (21) | 91.58 (20.3) | <0.001 | 92.65 (20.1) | 87.63 (18) | <0.001 |
| Respiratory rate, breaths/min | 22.17 (7.4) | 20.99 (7.4) | <0.001 | 22.05 (6.5) | 20.06 (5.9) | <0.001 |
| Temperature, C° | 36.74 (1) | 36.83 (0.8) | <0.001 | 36.78 (0.9) | 36.91 (0.8) | 0.047 |
| Systolic blood pressure, mmHg | 117.03 (26.6) | 119.69 (25.7) | <0.001 | 116.45 (25.3) | 113.48 (20.5) | 0.11 |
| **Blood count** |  |  |  |  |  |  |
| Hemoglobin, g/dL | 11.11 (2.4) | 11.35 (2.3) | <0.001 | 10.03 (2.1) | 10.48 (1.9) | 0.003 |
| Hematocrit, % | 34.02 (7.2) | 34.55 (6.9) | 0.004 | 30.83 (6.5) | 31.51 (5.4) | 0.15 |
| White blood cell count, 1000/μL | 13.45 (8.5) | 13.48 (7.7) | 0.9 | 14.54 (10.2) | 14.1 (7.3) | 0.55 |
| Platelet count, 1000/μL | 199.74 (100.9) | 206.7 (100.9) | 0.008 | 187.88 (115.9) | 184.88 (100.1) | 0.72 |
| MCV, fL | 91.45 (7.6) | 90.96 (6.9) | 0.014 | 92.66 (7.8) | 90.89 (6.5) | 0.002 |
| **Chemistry** |  |  |  |  |  |  |
| Na, mEq/L | 138.43 (5.6) | 138.54 (5.4) | 0.46 | 138.52 (5.2) | 138.62 (4.6) | 0.79 |
| K, mEq/L | 4.25 (0.8) | 4.16 (0.7) | <0.001 | 4.21 (0.6) | 4.29 (0.6) | 0.11 |
| Bicarbonate, mEq/L | 25.09 (8.1) | 24.24 (6.7) | <0.001 | 24.75 (6.9) | 23.26 (4.9) | 0.003 |
| Creatinine, mg/dL | 1.81 (1.6) | 1.54 (1.5) | <0.001 | 1.76 (1.6) | 1.44 (1.2) | 0.004 |
| Glucose, mg/dL | 157.14 (86.9) | 155.36 (74.7) | 0.42 | 159.41 (85.5) | 138.73 (55.7) | <0.001 |
| Lactate*, mmol/L | 3.04 (3.4) | 2.54 (2.6) | <0.001 | 2.37 (2.4) | 2.28 (2.1) | 0.61 |
| Total bilirubin*, mg/dL | 1.36 (3.2) | 1.21 (2.4) | 0.11 | 2.29 (5.5) | 2.07 (4.5) | 0.61 |
| **Other** |  |  |  |  |  |  |
| In-hospital mortality | 0.22 (0.4) | 0.15 (0.4) | <0.001 | 0.35 (0.5) | 0.18 (0.4) | <0.001 |

Patient characteristics are compared between White and Black patients in Supplementary Table 1. We confirmed that occult hypoxemia was more prevalent among Black patients (12.3% vs. 7.8% in eICU and 8.0% vs. 4.2% in MIMIC-IV). Of the clinical differences observed in occult hypoxemia cases, higher heart rate, higher respiratory rate, lower hemoglobin, lower MCV, higher lactate, and higher creatinine, were observed in Black patients compared to White patients. While there was no statistically significant difference in mortality by race in eICU, the in-hospital mortality was significantly higher among Black patients compared to White patients in MIMIC-IV (20% vs. 15%).

### Effect of race and covariates on occult hypoxemia

The unadjusted odds of having occult hypoxemia were significantly higher in Black vs. White individuals in both datasets (Odds ratio 1.66, [95% CI: 1.41, 1.95] in eICU and 2.00, [95% CI: 1.22, 3.14] in MIMIC-IV).

In the multivariable model (Figure 2, Supplementary Table 2), we found statistically significant associations of the odds of occult hypoxemia with younger age, higher PaCO2, lower arterial pH, higher respiratory rate, lower systolic blood pressure, lower hemoglobin level, and higher creatinine level in eICU. Similar patterns were observed in MIMIC-IV, although the differences in pH, systolic blood pressure did not reach statistical significance. After covariate adjustment, race was not statistically significantly associated with occult hypoxemia in both datasets. The proportion of the racial effect mediated by clinical factors included in this analysis was estimated to be 9.2% in eICU and 44.4% in MIMIC-IV.

**Figure 2.**
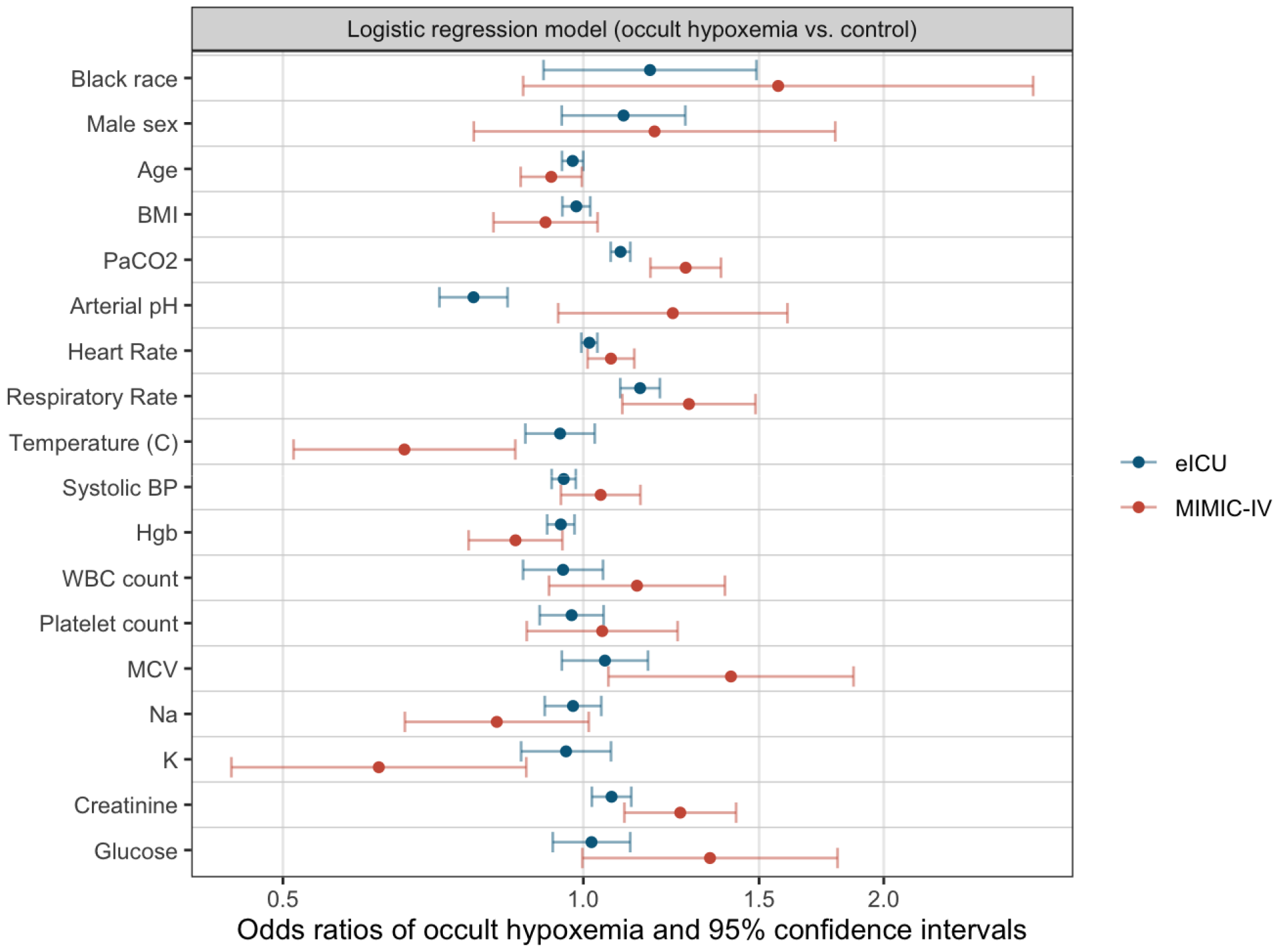
Odds ratios of occult hypoxemia for each covariate and the 95% confidence intervals in multivariable logistic regression models. Age, BMI, paCO2, heart rate, respiratory rate and sodium level are scaled by a factor of 5. Systolic blood pressure, white blood cell count and MCV are scaled by a factor of 10. Platelet count and glucose are scaled by a factor of 100. Arterial blood pH is scaled by a factor of 1/10.

All analyses were repeated using SaO2 and SpO2 pairs that are maximum 5 minutes apart and the results were consistent (Supplement C).

## Discussion

Despite the importance of commercially available pulse oximeters to rapidly identify hypoxemia, the reduced accuracy in Black patients is a major concern. Our study compared a comprehensive list of commonly available clinical markers between occult hypoxemia and control and confirmed some of the differences reported in previous studies (2, 4). We also compared differences in these clinical markers between Black and White individuals. We examined each effect in multivariable models and assessed how much of the racial effect on the prevalence of occult hypoxemia is mediated by the racial differences in these clinical markers.

In our study, Black patients were more likely to have clinical indicators associated with shock state or poor peripheral perfusion such as higher heart rate, respiratory rate, creatinine and lactate levels. Erickson et al. reported that Black patients tend to have more severe illness measured by Acute Physiology Score at the time of ICU admissions (19). Tolchin et al. found Black patients were more likely to have a higher SOFA score than White patients at the time of hospitalization (20). Our results suggest that Black patients may be sicker when they are treated in intensive care units, and racial disparities in the delivery of critical care medicine may be contributing to the racial difference in the prevalence of occult hypoxemia. Notably, a recent study by Gudelunas et al. suggested that low perfusion state is a significant contributor to the pulse oximetry errors when combined with darker skin pigmentation (21).

Our study has several limitations. While our study suggests certain clinical factors may account for racial differences in the prevalence of occult hypoxemia, our mediation analysis does not prove causal direction. We could not assess some important factors that may affect pulse oximetry accuracy due to data limitations, such skin tone, smoking status, COHb, MetHb, or use of nail colors. We did not include medication use such as vasopressors, as this could depend on provider practice or goals of care. We chose to not include comorbidities due to uncertainty about the accuracy of diagnosis codes, and model complexity.

## Conclusion

Clinical differences, in addition to skin tone, may play a role in the increased prevalence of occult hypoxemia in Black patients. Further studies are needed to assess whether accounting for these clinical discrepancies can reduce the racial differences in the prevalence of occult hypoxemia.

## Data Availability

The datasets analyzed during the current study are available under the links https://eicu-crd.mit.edu/ and https://mimic.mit.edu/. All data generated during this study are included in this published article and its supplementary information files.

## Declaration

### Ethics approval and consent to participate

The study is exempt from institutional review board approval due to the retrospective design, lack of direct patient intervention, and the security schema, for which the re-identification risk was certified as meeting safe harbor standards by an independent privacy expert (Privacert, Cambridge, MA) (Health Insurance Portability and Accountability Act Certification no. 1031219-2).

### Institutional Review Board (IRB) statement

Because our study did not fall under the board’s guidelines as human subjects research, no IRB review was necessary (and thus no number was assigned).

### Consent for publication

Not applicable.

### Authors’ contributions

SM and MC devised the research question and study design. SM curated, analyzed data, conducted literature review, and wrote the manuscript. NC and AJ conducted literature review and wrote the manuscript. CT reviewed the programming code for data analysis. NX and SE provided support for literature review. MS and MM provided statistical support. MM, BH and MC provided support for writing the manuscript and supervised the entire work. All authors read and approved the final manuscript.

---

**Supplementary Table 1.**
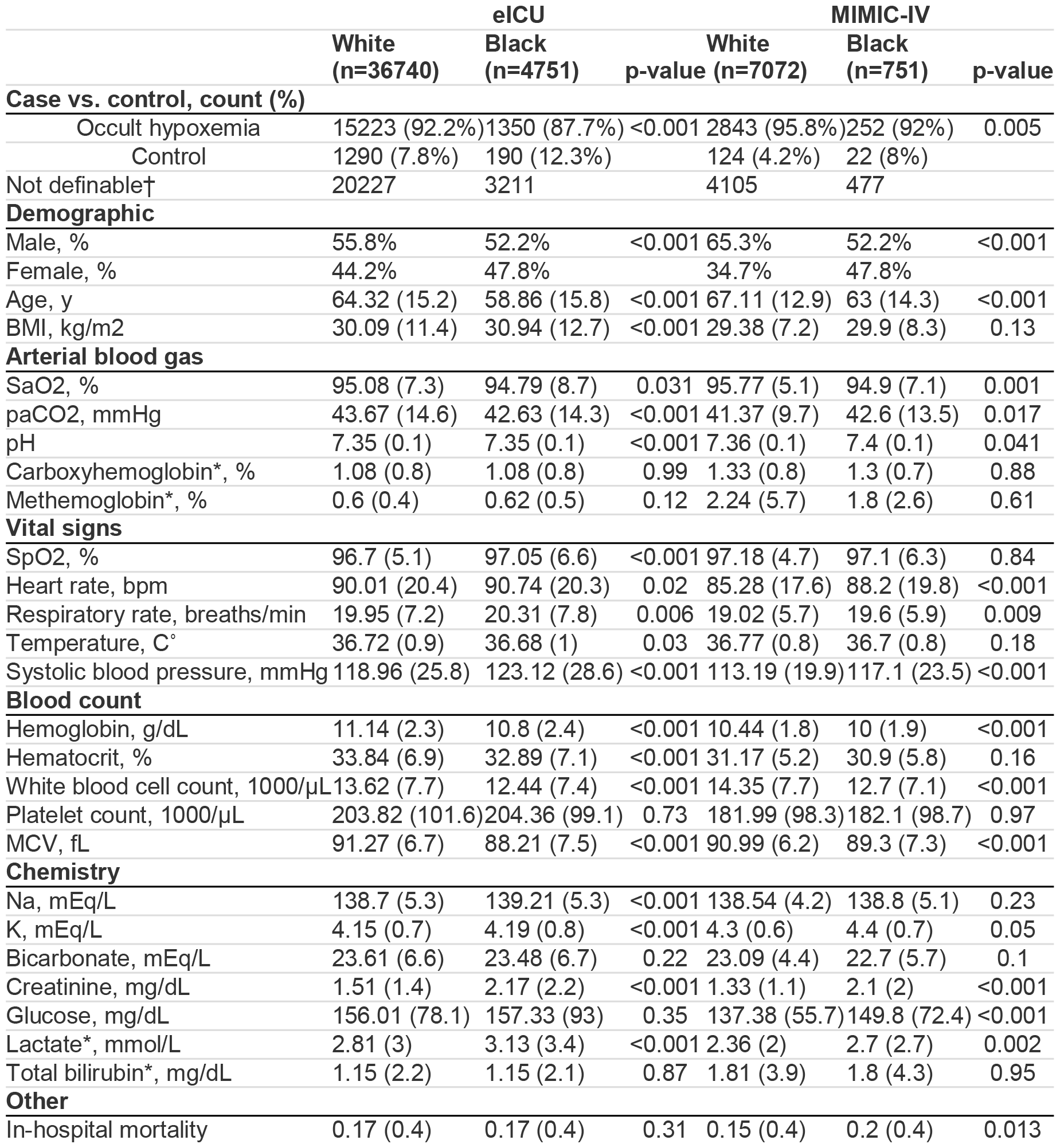
Patient characteristics by race. Univariable comparison between White and Black patients. †Occult hypoxemia could not be defined when SpO2 was not within 92 and 98%, or SaO2 was below 70%. *Missing values in more than 30% of observations.

**Supplementary Table 2.**
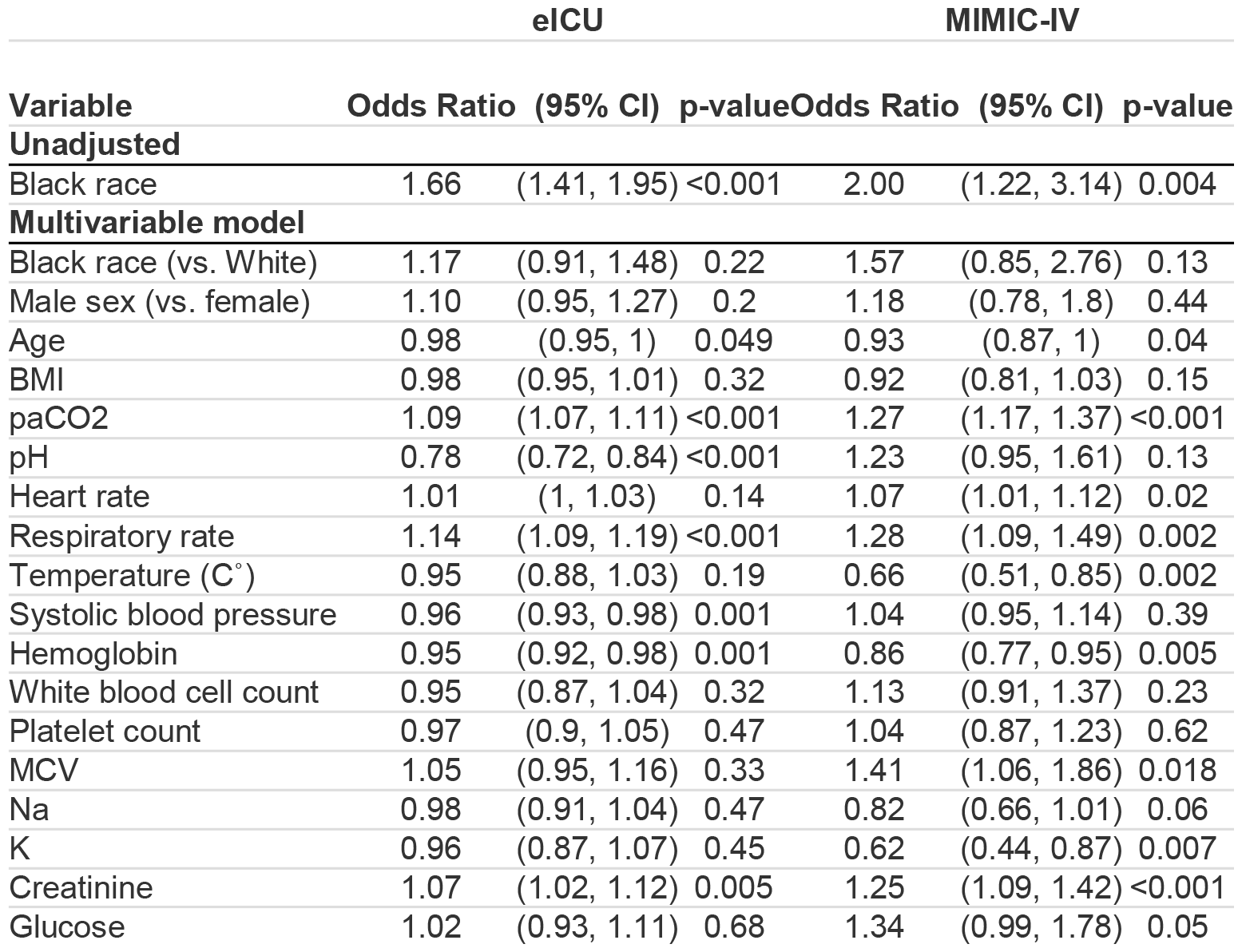
Odds ratios of occult hypoxemia in multivariable logistic regression models. Odds ratios, 95% confidence intervals and p-values of race and covariates are also shown. Age, BMI, paCO2, heart rate, respiratory rate and sodium level are scaled by a factor of 5. Systolic blood pressure, white blood cell count and MCV are scaled by a factor of 10. Platelet count and glucose are scaled by a factor of 100. Arterial blood pH is scaled by a factor of 1/10.

