## Supplement for "Clinical factors associated with racial differences in the prevalence of occult hypoxemia: a retrospective case-control study"

**Supplementary Material**

**Supplementary results**

**Supplementary Table 1. Patient characteristics by race** Univariable comparison between White and Black patients. †Occult hypoxemia could not be defined when SpO2 was not within 92 and 98%, or SaO2 was below 70%. *Missing values in more than 30% of observations.

|  | **eICU** | | | **MIMIC-IV** | | |
| --- | --- | --- | --- | --- | --- | --- |
|  | **White**  **(n=36740)** | **Black**  **(n=4751)** | **p-value** | **White**  **(n=7072)** | **Black**  **(n=751)** | **p-value** |
| **Case vs. control, count (%)** | | | | | | |
| Occult hypoxemia | 15223 (92.2%) | 1350 (87.7%) | <0.001 | 2843 (95.8%) | 252 (92%) | 0.005 |
| Control | 1290 (7.8%) | 190 (12.3%) |  | 124 (4.2%) | 22 (8%) |  |
| Not definable† | 20227 | 3211 |  | 4105 | 477 |  |
| **Demographic** | | | | | | |
| Male, % | 55.8% | 52.2% | <0.001 | 65.3% | 52.2% | <0.001 |
| Female, % | 44.2% | 47.8% |  | 34.7% | 47.8% |  |
| Age, y | 64.32 (15.2) | 58.86 (15.8) | <0.001 | 67.11 (12.9) | 63 (14.3) | <0.001 |
| BMI, kg/m2 | 30.09 (11.4) | 30.94 (12.7) | <0.001 | 29.38 (7.2) | 29.9 (8.3) | 0.13 |
| **Arterial blood gas** | | | | | | |
| SaO2, % | 95.08 (7.3) | 94.79 (8.7) | 0.031 | 95.77 (5.1) | 94.9 (7.1) | 0.001 |
| paCO2, mmHg | 43.67 (14.6) | 42.63 (14.3) | <0.001 | 41.37 (9.7) | 42.6 (13.5) | 0.017 |
| pH | 7.35 (0.1) | 7.35 (0.1) | <0.001 | 7.36 (0.1) | 7.4 (0.1) | 0.041 |
| Carboxyhemoglobin*, % | 1.08 (0.8) | 1.08 (0.8) | 0.99 | 1.33 (0.8) | 1.3 (0.7) | 0.88 |
| Methemoglobin*, % | 0.6 (0.4) | 0.62 (0.5) | 0.12 | 2.24 (5.7) | 1.8 (2.6) | 0.61 |
| **Vital signs** | | | | | | |
| SpO2, % | 96.7 (5.1) | 97.05 (6.6) | <0.001 | 97.18 (4.7) | 97.1 (6.3) | 0.84 |
| Heart rate, bpm | 90.01 (20.4) | 90.74 (20.3) | 0.02 | 85.28 (17.6) | 88.2 (19.8) | <0.001 |
| Respiratory rate, breaths/min | 19.95 (7.2) | 20.31 (7.8) | 0.006 | 19.02 (5.7) | 19.6 (5.9) | 0.009 |
| Temperature, C˚ | 36.72 (0.9) | 36.68 (1) | 0.03 | 36.77 (0.8) | 36.7 (0.8) | 0.18 |
| Systolic blood pressure, mmHg | 118.96 (25.8) | 123.12 (28.6) | <0.001 | 113.19 (19.9) | 117.1 (23.5) | <0.001 |
| **Blood count** | | | | | | |
| Hemoglobin, g/dL | 11.14 (2.3) | 10.8 (2.4) | <0.001 | 10.44 (1.8) | 10 (1.9) | <0.001 |
| Hematocrit, % | 33.84 (6.9) | 32.89 (7.1) | <0.001 | 31.17 (5.2) | 30.9 (5.8) | 0.16 |
| White blood cell count, 1000/µL | 13.62 (7.7) | 12.44 (7.4) | <0.001 | 14.35 (7.7) | 12.7 (7.1) | <0.001 |
| Platelet count, 1000/µL | 203.82 (101.6) | 204.36 (99.1) | 0.73 | 181.99 (98.3) | 182.1 (98.7) | 0.97 |
| MCV, fL | 91.27 (6.7) | 88.21 (7.5) | <0.001 | 90.99 (6.2) | 89.3 (7.3) | <0.001 |
| **Chemistry** | | | | | | |
| Na, mEq/L | 138.7 (5.3) | 139.21 (5.3) | <0.001 | 138.54 (4.2) | 138.8 (5.1) | 0.23 |
| K, mEq/L | 4.15 (0.7) | 4.19 (0.8) | <0.001 | 4.3 (0.6) | 4.4 (0.7) | 0.05 |
| Bicarbonate, mEq/L | 23.61 (6.6) | 23.48 (6.7) | 0.22 | 23.09 (4.4) | 22.7 (5.7) | 0.1 |
| Creatinine, mg/dL | 1.51 (1.4) | 2.17 (2.2) | <0.001 | 1.33 (1.1) | 2.1 (2) | <0.001 |
| Glucose, mg/dL | 156.01 (78.1) | 157.33 (93) | 0.35 | 137.38 (55.7) | 149.8 (72.4) | <0.001 |
| Lactate*, mmol/L | 2.81 (3) | 3.13 (3.4) | <0.001 | 2.36 (2) | 2.7 (2.7) | 0.002 |
| Total bilirubin*, mg/dL | 1.15 (2.2) | 1.15 (2.1) | 0.87 | 1.81 (3.9) | 1.8 (4.3) | 0.95 |
| **Other** | | | | | | |
| In-hospital mortality | 0.17 (0.4) | 0.17 (0.4) | 0.31 | 0.15 (0.4) | 0.2 (0.4) | 0.013 |

**Supplementary Table 2. Odds ratios of occult hypoxemia in multivariable logistic regression models.** Odds ratios, 95% confidence intervals and p-values of race and covariates are also shown. Age, BMI, paCO2, heart rate, respiratory rate and sodium level are scaled by a factor of 5. Systolic blood pressure, white blood cell count and MCV are scaled by a factor of 10. Platelet count and glucose are scaled by a factor of 100. Arterial blood pH is scaled by a factor of 1/10.

|  | **eICU** | | | **MIMIC-IV** | | |
| --- | --- | --- | --- | --- | --- | --- |
| **Variable** | **Odds Ratio** | **(95% CI)** | **p-value** | **Odds Ratio** | **(95% CI)** | **p-value** |
| **Unadjusted** | | | | | | |
| Black race | 1.66 | (1.41, 1.95) | <0.001 | 2.00 | (1.22, 3.14) | 0.004 |
| **Multivariable model** | | | | | | |
| Black race (vs. White) | 1.17 | (0.91, 1.48) | 0.22 | 1.57 | (0.85, 2.76) | 0.13 |
| Male sex (vs. female) | 1.10 | (0.95, 1.27) | 0.2 | 1.18 | (0.78, 1.8) | 0.44 |
| Age | 0.98 | (0.95, 1) | 0.049 | 0.93 | (0.87, 1) | 0.04 |
| BMI | 0.98 | (0.95, 1.01) | 0.32 | 0.92 | (0.81, 1.03) | 0.15 |
| paCO2 | 1.09 | (1.07, 1.11) | <0.001 | 1.27 | (1.17, 1.37) | <0.001 |
| pH | 0.78 | (0.72, 0.84) | <0.001 | 1.23 | (0.95, 1.61) | 0.13 |
| Heart rate | 1.01 | (1, 1.03) | 0.14 | 1.07 | (1.01, 1.12) | 0.02 |
| Respiratory rate | 1.14 | (1.09, 1.19) | <0.001 | 1.28 | (1.09, 1.49) | 0.002 |
| Temperature (C˚) | 0.95 | (0.88, 1.03) | 0.19 | 0.66 | (0.51, 0.85) | 0.002 |
| Systolic blood pressure | 0.96 | (0.93, 0.98) | 0.001 | 1.04 | (0.95, 1.14) | 0.39 |
| Hemoglobin | 0.95 | (0.92, 0.98) | 0.001 | 0.86 | (0.77, 0.95) | 0.005 |
| White blood cell count | 0.95 | (0.87, 1.04) | 0.32 | 1.13 | (0.91, 1.37) | 0.23 |
| Platelet count | 0.97 | (0.9, 1.05) | 0.47 | 1.04 | (0.87, 1.23) | 0.62 |
| MCV | 1.05 | (0.95, 1.16) | 0.33 | 1.41 | (1.06, 1.86) | 0.018 |
| Na | 0.98 | (0.91, 1.04) | 0.47 | 0.82 | (0.66, 1.01) | 0.06 |
| K | 0.96 | (0.87, 1.07) | 0.45 | 0.62 | (0.44, 0.87) | 0.007 |
| Creatinine | 1.07 | (1.02, 1.12) | 0.005 | 1.25 | (1.09, 1.42) | <0.001 |
| Glucose | 1.02 | (0.93, 1.11) | 0.68 | 1.34 | (0.99, 1.78) | 0.05 |

### Supplement A. DAG considered in the study design

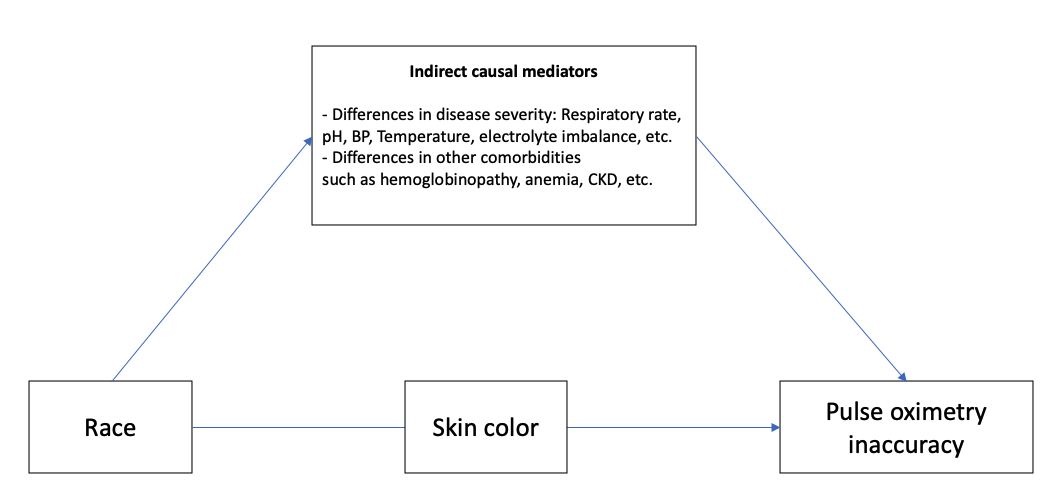

While some factors may also be influenced by the outcome, i.e., pulse oximetry inaccuracy, it is difficult to determine whether these are causes or results of the outcome. E.g., respiratory rate, or pH. One of the limitations of our study is that the causal directions cannot be proven.

**Supplement B. Outlier removal, ranges, quantiles, and histograms of extracted data**

##### **Outlier removal**

In our analysis of the data, we noted several outliers, such as 9999 in lab data, height below 50 cm or temperature above 50 C˚ or below 30 F˚. These outliers raised two concerns; first, in many cases it was difficult to determine whether a value is a true extreme or a data entry error, and second, for statistical analyses, extreme values can violate statistical assumptions. Most studies utilizing eICU or MIMIC data do not report how outliers were handled (13, 21). To address the problem of outliers, we tested three different methods to deal with outliers; removing 1) 0.1% at each end, 2) 0.1% at each end and values 6 standard deviations beyond the mean, 3) 0.1% at each end and values 4 standard deviations beyond the mean. We used the results using the least restrictive outlier removal method, removing 0.1% at each end, as our primary analysis, and used the other methods to examine the sensitivity of our results to varying outlier removal algorithms. We confirmed that the results are consistent with all three outlier removal methods. To minimize data loss, we applied the outlier removal method to each variable separately prior to applying the inclusion criteria and constructing the final dataset.

**Ranges, quantiles, and histograms of data after removing outliers beyond 0.1% at each end**

#### B.1. Data ranges and quantiles - eICU

| **Variable name** | **min** | **25%** | **50%** | **75%** | **max** |
| --- | --- | --- | --- | --- | --- |
| Age | 18.00 | 54.00 | 65.00 | 75.00 | 89.00 |
| BMI | 11.69 | 23.79 | 28.11 | 33.74 | 378.79 |
| Heart rate | 39.00 | 75.00 | 88.00 | 103.00 | 164.00 |
| Respiratory rate | 1.00 | 15.00 | 19.00 | 24.00 | 74.00 |
| Temperature (C˚) | 31.70 | 36.30 | 36.70 | 37.20 | 40.28 |
| Systolic blood pressure | 3.00 | 101.00 | 116.00 | 135.00 | 291.00 |
| pH | 6.83 | 7.30 | 7.36 | 7.42 | 7.65 |
| HCO3 | 3.50 | 20.00 | 23.00 | 26.50 | 54.50 |
| paCO2 | 12.10 | 34.00 | 40.50 | 48.30 | 128.00 |
| Hemoglobin | 4.60 | 9.40 | 11.00 | 12.70 | 18.20 |
| Hematocrit | 14.10 | 28.60 | 33.30 | 38.30 | 54.90 |
| White blood cell count | 0.11 | 8.47 | 11.90 | 16.60 | 83.00 |
| Platelet count | 7.00 | 135.00 | 188.00 | 253.00 | 927.00 |
| MCV | 62.80 | 86.80 | 90.80 | 95.00 | 117.70 |
| Carboxyhemoglobin | 0.10 | 0.40 | 0.90 | 1.40 | 9.10 |
| Methemoglobin | 0.10 | 0.30 | 0.40 | 0.70 | 3.60 |
| Na | 113.00 | 136.00 | 139.00 | 142.00 | 166.00 |
| K | 2.30 | 3.70 | 4.10 | 4.50 | 7.40 |
| Glucose | 38.00 | 110.00 | 137.00 | 177.00 | 907.00 |
| Creatinine | 0.20 | 0.77 | 1.07 | 1.72 | 14.49 |
| Lactate | 0.31 | 1.10 | 1.80 | 3.20 | 24.80 |
| Total bilirubin | 0.13 | 0.40 | 0.60 | 1.10 | 39.60 |

#### B.2. Histograms -eICU

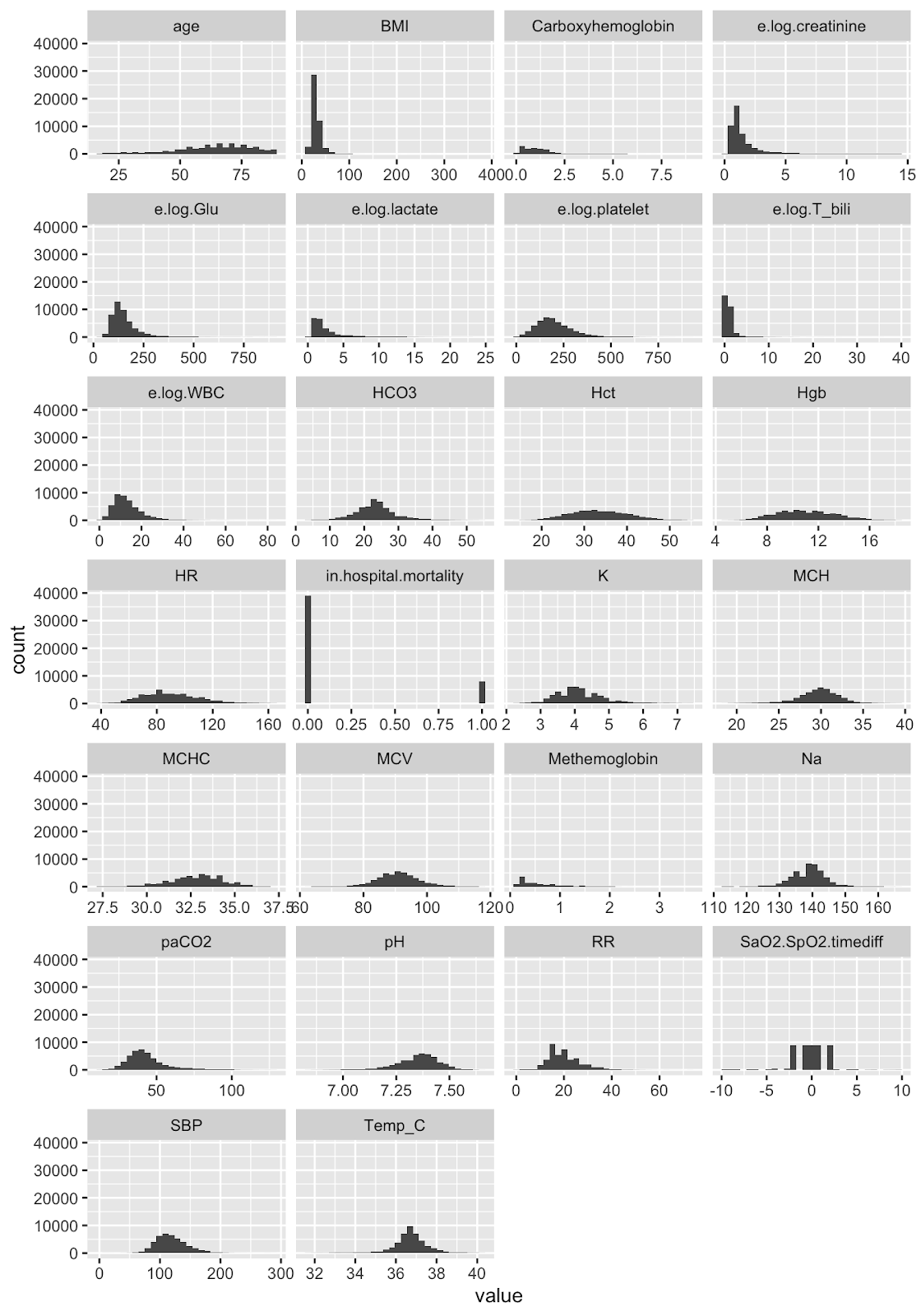

#### B.3. Data ranges and quantiles MIMIC

| **Variable name** | **min** | **25%** | **50%** | **75%** | **max** |
| --- | --- | --- | --- | --- | --- |
| Age | 18.00 | 58.00 | 67.00 | 76.00 | 89.0 |
| BMI | 11.89 | 24.59 | 28.12 | 32.65 | 114.5 |
| Heart rate | 39.00 | 74.00 | 83.00 | 96.00 | 160.0 |
| Respiratory rate | 2.00 | 15.00 | 18.00 | 22.00 | 47.0 |
| Temperature (C˚) | 32.70 | 36.39 | 36.72 | 37.17 | 39.9 |
| Systolic blood pressure | 44.00 | 100.00 | 111.00 | 125.00 | 209.0 |
| pH | 6.94 | 7.32 | 7.37 | 7.41 | 7.6 |
| HCO3 | 8.00 | 21.00 | 23.00 | 25.00 | 46.0 |
| paCO2 | 18.00 | 36.00 | 40.00 | 45.00 | 109.0 |
| Hemoglobin | 4.90 | 9.10 | 10.30 | 11.60 | 17.2 |
| Hematocrit | 15.50 | 27.40 | 30.60 | 34.60 | 52.2 |
| White blood cell count | 0.20 | 9.40 | 12.80 | 17.40 | 94.9 |
| Platelet count | 9.00 | 121.00 | 161.00 | 217.00 | 994.0 |
| MCV | 68.00 | 87.00 | 90.00 | 94.00 | 121.0 |
| Carboxyhemoglobin | 0.10 | 1.00 | 1.00 | 1.70 | 6.0 |
| Methemoglobin | 0.10 | 0.30 | 1.00 | 1.00 | 35.0 |
| Na | 119.00 | 136.00 | 139.00 | 141.00 | 162.0 |
| K | 2.50 | 3.90 | 4.30 | 4.70 | 7.8 |
| Glucose | 41.00 | 107.00 | 125.00 | 154.00 | 669.0 |
| Creatinine | 0.30 | 0.70 | 1.00 | 1.50 | 11.4 |
| Lactate | 0.60 | 1.30 | 1.80 | 2.80 | 21.2 |
| Total bilirubin | 0.20 | 0.40 | 0.70 | 1.50 | 55.0 |

#### B.4. Histograms -MIMIC

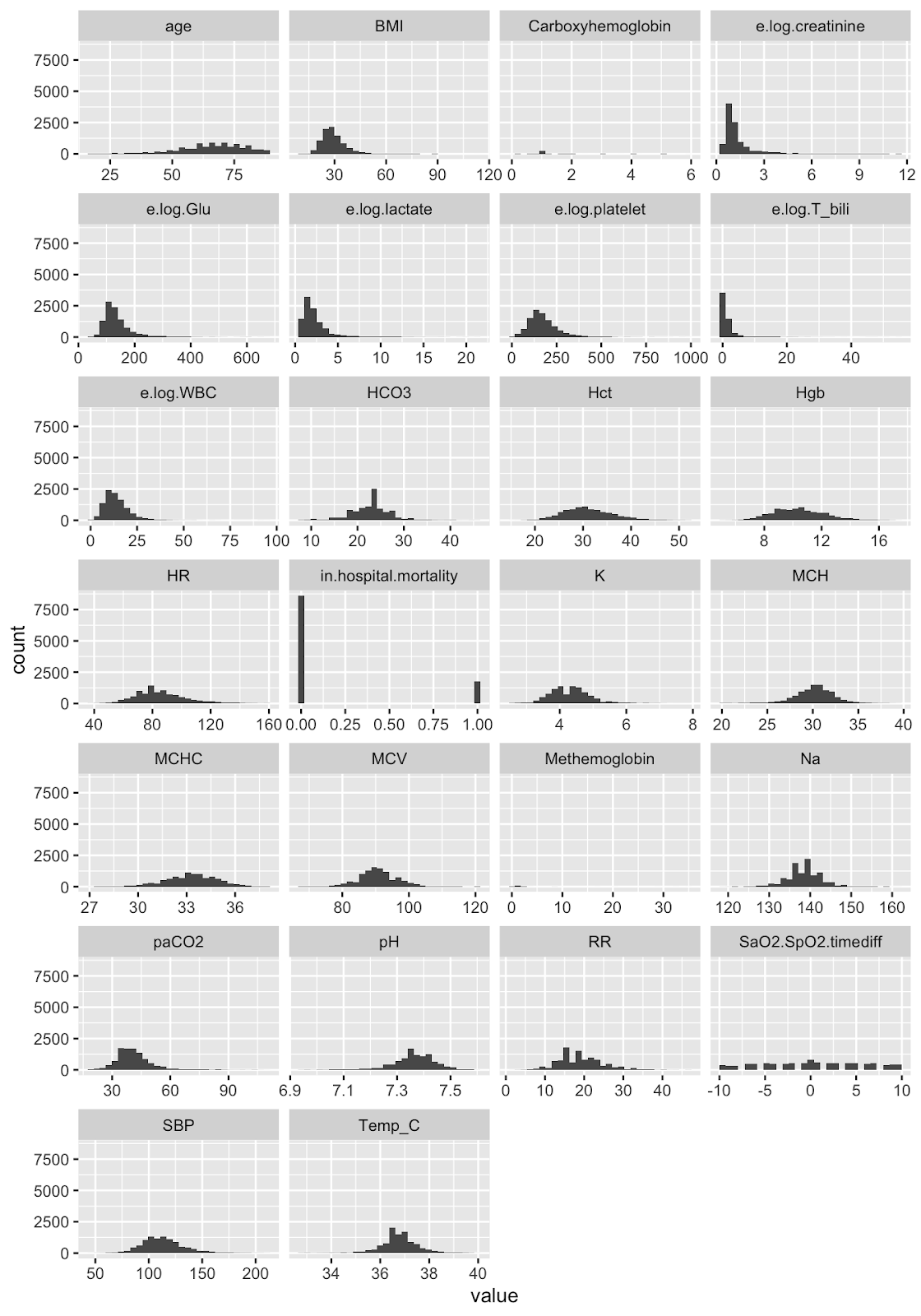

**Supplement C.** SaO2 and SpO2 pairs maximum 5 minutes apart

**5min Table1. Patient characteristics by occult hypoxemia vs. control** Univariable comparison between cases and controls. *Missing values in more than 30% of observations.

|  | **eICU** | | | **MIMIC-IV** | | |
| --- | --- | --- | --- | --- | --- | --- |
|  | **Occult hypoxemia (n=1614)** | **Control (n=18162)** | **p-value** | **Occult hypoxemia (n=117)** | **Control (n=2359)** | **p-value** |
| **Demographic** | | | | | | |
| **Race/Ethnicity, %** | | | | | | |
| White | 78.1% | 81.7% | <0.001 | 60.7% | 68.9% | 0.12 |
| Black | 11.2% | 7.2% |  | 12% | 6.4% |  |
| Hispanic/Latino | 4.5% | 5.6% |  | 20.5% | 18.7% |  |
| Asian | 4.2% | 3.7% |  | 6% | 3.8% |  |
| American Indian | 1.4% | 1.1% |  | 0.9% | 2.2% |  |
| Other/Unknown | 0.7% | 0.7% |  |  | 0.1% |  |
| Male, % | 55.8% | 56% | 0.93 | 60.7% | 62.2% | 0.82 |
| Female, % | 44.2% | 44% |  | 39.3% | 37.8% |  |
| Age, y | 63.36 (14.5) | 64.03 (15.1) | 0.08 | 62.77 (13.7) | 65.88 (13.8) | 0.018 |
| BMI, kg/m2 | 31.5 (14) | 31.13 (11.6) | 0.32 | 30.36 (8.9) | 30.17 (7.8) | 0.84 |
| **Arterial blood gas** | | | | | | |
| SaO2, % | 85.37 (4.7) | 95.45 (2.5) | <0.001 | 85.62 (4.7) | 95.61 (2.1) | <0.001 |
| paCO2, mmHg | 50.85 (18.5) | 44.34 (15) | <0.001 | 48.32 (15.8) | 42.08 (10.5) | <0.001 |
| pH | 7.31 (0.1) | 7.36 (0.1) | <0.001 | 7.35 (0.1) | 7.36 (0.1) | 0.37 |
| Carboxyhemoglobin*, % | 1.37 (1) | 1.14 (0.9) | <0.001 | 1.21 (0.6) | 1.24 (0.7) | 0.92 |
| Methemoglobin*, % | 0.63 (0.4) | 0.54 (0.4) | <0.001 | 0.45 (0.5) | 1.86 (4) | 0.08 |
| **Vital signs** | | | | | | |
| SpO2, % | 94.45 (1.9) | 95.99 (1.8) | <0.001 | 94.52 (1.9) | 95.97 (1.8) | <0.001 |
| Heart rate, bpm | 93.86 (21) | 91.55 (20.3) | <0.001 | 92.89 (20.7) | 87.98 (18.1) | 0.013 |
| Respiratory rate, breaths/min | 22.2 (7.4) | 20.99 (7.4) | <0.001 | 22 (6.8) | 20.07 (6) | 0.003 |
| Temperature, C˚ | 36.75 (1) | 36.83 (0.8) | <0.001 | 36.87 (0.8) | 36.9 (0.8) | 0.69 |
| Systolic blood pressure, mmHg | 117.1 (26.7) | 119.7 (25.7) | <0.001 | 116.64 (24.6) | 113.61 (20.6) | 0.19 |
| **Blood count** | | | | | | |
| Hemoglobin, g/dL | 11.11 (2.4) | 11.36 (2.3) | <0.001 | 9.94 (1.9) | 10.44 (1.9) | 0.007 |
| Hematocrit, % | 34 (7.2) | 34.56 (6.9) | 0.003 | 30.5 (6.1) | 31.41 (5.4) | 0.12 |
| White blood cell count, 1000/µL | 13.45 (8.5) | 13.47 (7.7) | 0.92 | 14.49 (10.7) | 14.17 (7.4) | 0.75 |
| Platelet count, 1000/µL | 199.61 (101.1) | 206.94 (101.1) | 0.006 | 192.78 (119.7) | 184.69 (97.2) | 0.47 |
| MCV, fL | 91.47 (7.6) | 90.97 (6.9) | 0.013 | 92.27 (7.7) | 90.8 (6.5) | 0.046 |
| **Chemistry** | | | | | | |
| Na, mEq/L | 138.39 (5.6) | 138.53 (5.4) | 0.34 | 138.13 (5.1) | 138.62 (4.6) | 0.31 |
| K, mEq/L | 4.25 (0.8) | 4.16 (0.7) | <0.001 | 4.26 (0.6) | 4.28 (0.6) | 0.77 |
| Bicarbonate, mEq/L | 25.14 (8.1) | 24.25 (6.7) | <0.001 | 24.62 (6.1) | 23.2 (4.8) | 0.013 |
| Creatinine, mg/dL | 1.81 (1.6) | 1.54 (1.5) | <0.001 | 1.88 (1.7) | 1.42 (1.2) | 0.004 |
| Glucose, mg/dL | 156.87 (87) | 155.38 (74.8) | 0.51 | 168.67 (91.2) | 138.35 (52.1) | <0.001 |
| Lactate*, mmol/L | 3.02 (3.4) | 2.53 (2.5) | <0.001 | 2.5 (2.6) | 2.36 (2.1) | 0.56 |
| Total bilirubin*, mg/dL | 1.35 (3.1) | 1.21 (2.4) | 0.15 | 2.48 (6.4) | 2.1 (4.5) | 0.56 |
| **Other** | | | | | | |
| In-hospital mortality | 0.22 (0.4) | 0.15 (0.4) | <0.001 | 0.31 (0.5) | 0.19 (0.4) | 0.007 |

**5min Figure 1. Odds ratios of occult hypoxemia for each covariate and the 95% confidence intervals in multivariable logistic regression models.** Age, BMI, paCO2, heart rate, respiratory rate and sodium level are scaled by a factor of 5. Systolic blood pressure, white blood cell count and MCV are scaled by a factor of 10. Platelet count and glucose are scaled by a factor of 100. Arterial blood pH is scaled by a factor of 1/10.

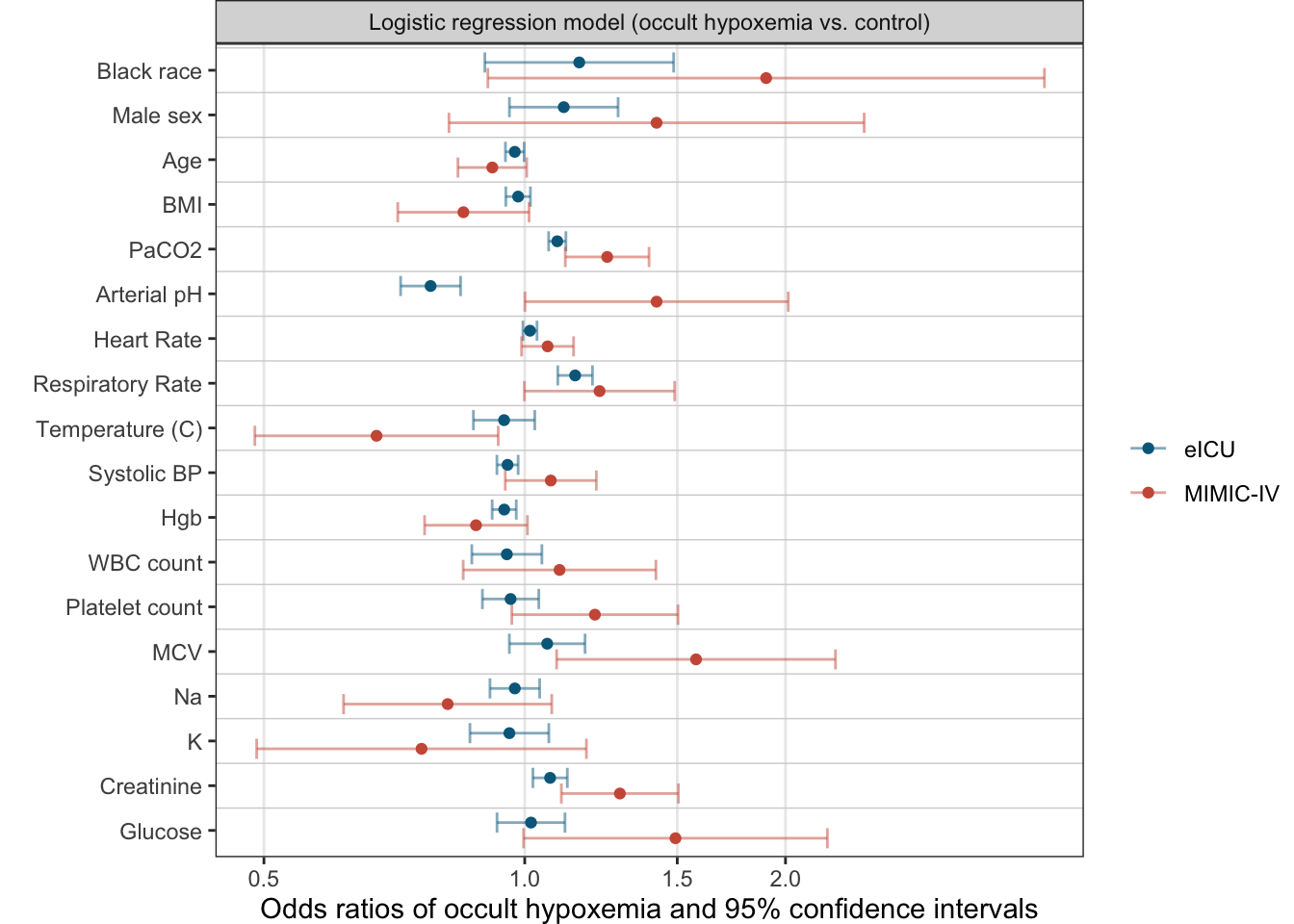

**Mediation analysis**

The mediated proportion of racial effect by other clinical factors is

eICU: 5.2%

MIMIC-IV: 32.1%.
